# Shifting patterns of importation risk of Bundibugyo Ebola virus disease to Europe under outbreak expansion scenarios

**DOI:** 10.64898/2026.05.31.26354511

**Authors:** Federico Fanelli, Francesco Parino, Chiara Poletto, Vittoria Colizza

**Author notes:** Co-first author (Federico Fanelli, Francesco Parino).

## Abstract

The 2026 Bundibugyo Ebola outbreak in eastern Democratic Republic of the Congo (DRC) has already generated international spread to Uganda, raising concerns about further regional and international dissemination. Using International Air Transport Association origin–destination passenger flows, we assessed relative exposure to Ebola virus disease importation into Europe under six outbreak expansion scenarios reflecting plausible pathways of geographical spread, including cross-border transmission and amplification in highly connected regional capitals. Relative exposure patterns remained largely unchanged under localized transmission in eastern DRC and border-spillover scenarios. Expansion into South Sudan generated a first structural increase in importation pressure to Europe through the connectivity associated with Juba, while hypothetical amplification in Kampala, Kigali, and Kinshasa substantially increased importation pressure and reshaped exposure patterns across Europe. Across all scenarios, France, Italy, and the United Kingdom remained among the most exposed countries. Mobility-informed scenario analyses support preparedness as the geography of the outbreak evolves.

## Text

As of 29 May 2026, the Bundibugyo Ebola outbreak, a Public Health Emergency of International Concern^1^, involved 125 confirmed cases (17 deaths), and 906 suspected cases (223 deaths) across Ituri, North Kivu, and South Kivu provinces in Eastern DRC, and 9 confirmed cases (1 death) in Uganda^2^.

Here, we assessed the relative risk of Ebola virus disease importation^3^ into Europe through commercial air travel, using International Air Transport Association origin–destination passenger flows, including connected itineraries. We considered six outbreak expansion scenarios designed to reflect plausible pathways of geographical spread from the currently affected area through porous borders (**Fig. 1a, Appendix**). Scenario 1 considered the currently reported geographical configuration of the country in Eastern DRC, including airports serving the provinces of Ituri, North Kivu, and South Kivu. Scenario 2 considered potential spillover to Western Uganda due to cross-border mobility, and included Ugandan airports close to the DRC border. Scenario 3 further considered cross-border transmission to South Sudan, with the inclusion of Juba, the country capital and main gateway of the neighboring country. Scenarios 4–6 explored hypothetical amplification assuming community transmission in major urban hubs—Kampala, Kigali, and Kinshasa.

**Figure 1.**
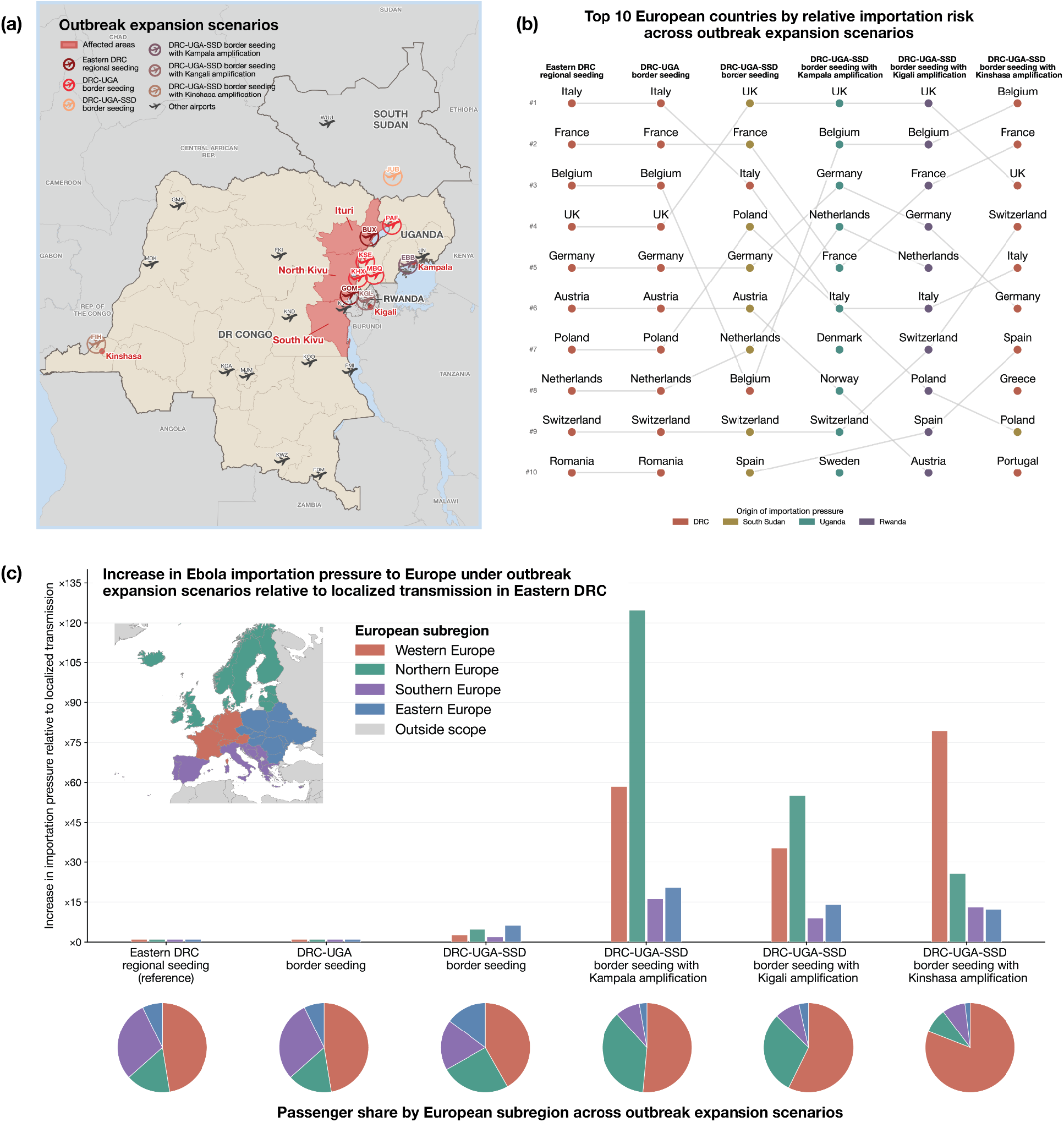
Ebola virus importation risk to Europe under six outbreak expansion scenarios. (a) Map of outbreak expansion scenarios corresponding to the seedings considered in the analysis: (i) eastern DRC regional seeding (Ituri, North Kivu, and South Kivu), (ii) DRC–Uganda border seeding, (iii) DRC–Uganda–South Sudan border seeding, and three hypothetical amplification scenarios involving (iv) Kampala (Uganda), (v) Kigali (Rwanda), and (vi) Kinshasa (DRC). Colored airport symbols indicate airports included as seed locations in each scenario. (b) Top 10 European countries by relative importation risk across scenarios. Dot colors indicate the dominant origin of importation pressure (DRC, Uganda, South Sudan, or Rwanda). The full ranking is provided in the **Appendix**. (c) Increase in Ebola importation pressure to European subregions under outbreak expansion scenarios, compared with localized transmission in eastern DRC (reference scenario). Pie charts indicate the distribution of passenger flows across subregions for each scenario.

Potential spread across the Ugandan border produced no change in either the relative exposure of European countries (**Fig. 1b**) or their overall importation pressure (**Fig. 1c**), compared with the currently reported localized transmission in eastern DRC, with Italy, France, Belgium, and the UK being the most exposed. Expansion across the border to South Sudan produced a first structural increase in importation pressure through the connectivity associated to Juba (5-fold increase for Northern Europe and 6-fold increase for Eastern Europe relative to localized transmission in DRC), while still leaving UK, France, and Italy among the highest-risk countries. Further amplification in highly connected capitals in the region substantially altered importation patterns. Kampala and Kigali amplification increased exposure in Northern Europe, with Kampala producing a >120-fold increase in importation pressure relative to localized transmission in DRC. Kinshasa amplification instead primarily reinforced exposure in Western Europe (~80-fold increase). In each case, the inclusion of the capital became the dominant source of importation pressure, reflecting its strong international connectivity.

Patterns of Ebola virus importation risk to Europe may rapidly shift under uncertainty surrounding the geographical extent and evolution of the outbreak, particularly if transmission expands into major urban hubs with strong international connectivity near the affected area. Although the current ECDC assessment considers the risk for the general population in the EU/EEA to be very low^4^, preparedness remains essential given the potential increase in importation pressure under amplification scenarios. In this context, WHO recommendations on cross-border screening, exit screening, and restriction of travel for contacts and confirmed cases should be prioritized. By contrast, broad travel restrictions or border closures are expected to provide only limited delays in international spread and may be counterproductive by hindering resource mobilization and response operations^5^. Strengthening surveillance, diagnostics, isolation, and response capacities in affected areas remains essential to limit further geographical spread.

## Data Availability

The global flight network data are commercially available from the International Air Transport Association (IATA, https://www.iata.org/en/contact-support).

## Acknowledgments

This study was partially supported by: the EU Horizon Europe grant ESCAPE (101095619) to F.P, V.C.; the ANRS Maladies Infectieuses Émergentes and France 2030/SGPI grant PReViX (ANRS-24-PEPRMIE-0003) to V.C.; the ANRS Maladies Infectieuses Émergentes ESCAPE project (Start Programme - Fellowships funding scheme - PhD grants 2025 ANRS00850-R-AR1) to F.F., V.C.. All authors acknowledge the Working Group on Pandemic Preparedness of the French Research Action on Modeling Epidemics (FRAME, AC Modélisation des maladies infectieuses) funded by the ANRS Maladies Infectieuses Émergentes.

## APPENDIX

### Air travel data

We use origin–destination (O&D) passenger flows derived from the 2024 International Air Transport Association (IATA) database to reconstruct intercontinental and regional travel patterns at the airport level. The IATA O&D data capture complete passenger itineraries, including traffic travelling via connecting flights rather than only direct flight segments. All analyses presented in this report are based on the cumulative passenger traffic reported for May, June, and July 2024. Data from 2024 showed moderate seasonal variability in passenger traffic to Europe. Relative to the annual traffic to Europe, mean passenger volumes during May– July ranged from 3% below to 16% above the monthly 2024 average across scenarios.

### Outbreak expansion scenarios

### Estimation of the relative risk of importation

The relative importation risk to country *c* was estimated as the proportion of total outgoing passenger flow from the selected seed airports directed to the country, for each scenario:

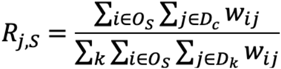

where *w*_*ij*_ denotes the origin–destination passenger flow from airport *i*, among the set *O*_*S*_ of origin airports included in scenario *S*, and airport j in the set *D*_*c*_ of airports of the destination destination country *c*.

The relative risk of importation for each European country was normalized to the total passenger flow from the source airports to Europe. Risks were then stratified by subregion [1]. For this study, Europe was defined according to the definition used in previous works on early-stage importation risk to Europe during the COVID-19 outbreak [2].

**Figure S1** reports the full list of ranked European countries under the six outbreak expansion scenarios (whereas the main text reports only the Top 10).

**Figure S1.**
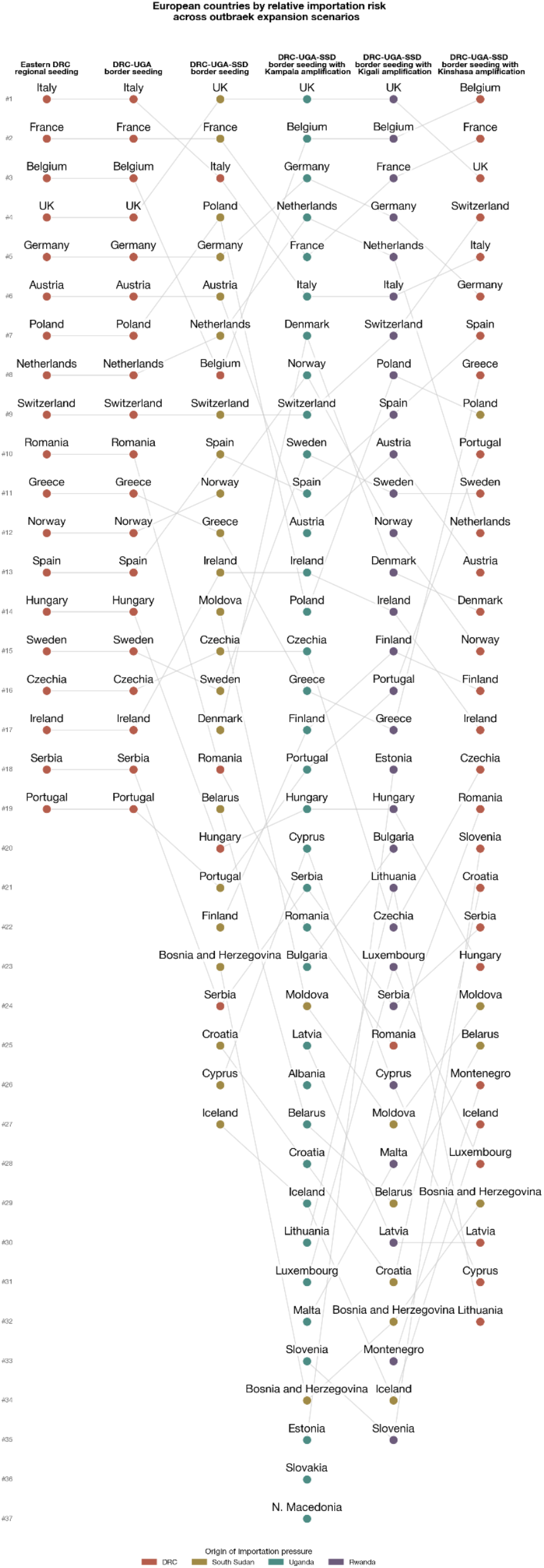
Ebola virus importation risk to Europe. Ranking of European countries by relative importation risk across outbreak expansion scenarios. Dot colors indicate the dominant origin of importation pressure (DRC, Uganda, South Sudan, or Rwanda). Analyses performed using the May-July 2024 traffic data.

### Estimation of the importation pressure

For each destination country, we estimated the origin importation pressure as the source country with the largest share of incoming international passenger traffic to the country.

The increase in importation pressure for each European subregion was estimated as the relative increase in importation risk under each outbreak expansion scenario compared with the localized eastern DRC transmission scenario, and computed from outbound origin-destination trips from the affected area.

### Sensitivity analysis on period of analysis

To account for potential seasonal effects in travel patterns, we conducted the analysis using average monthly data from the full 2024. Results do not show notable differences with those reported in the main text (**Figure S2, S3**), with the exception of the Kampala amplification scenario, in which the increase in importation pressure for Northern Europe rises to approximately 190-fold when using monthly averages of 2024 travel data.

**Figure S2.**
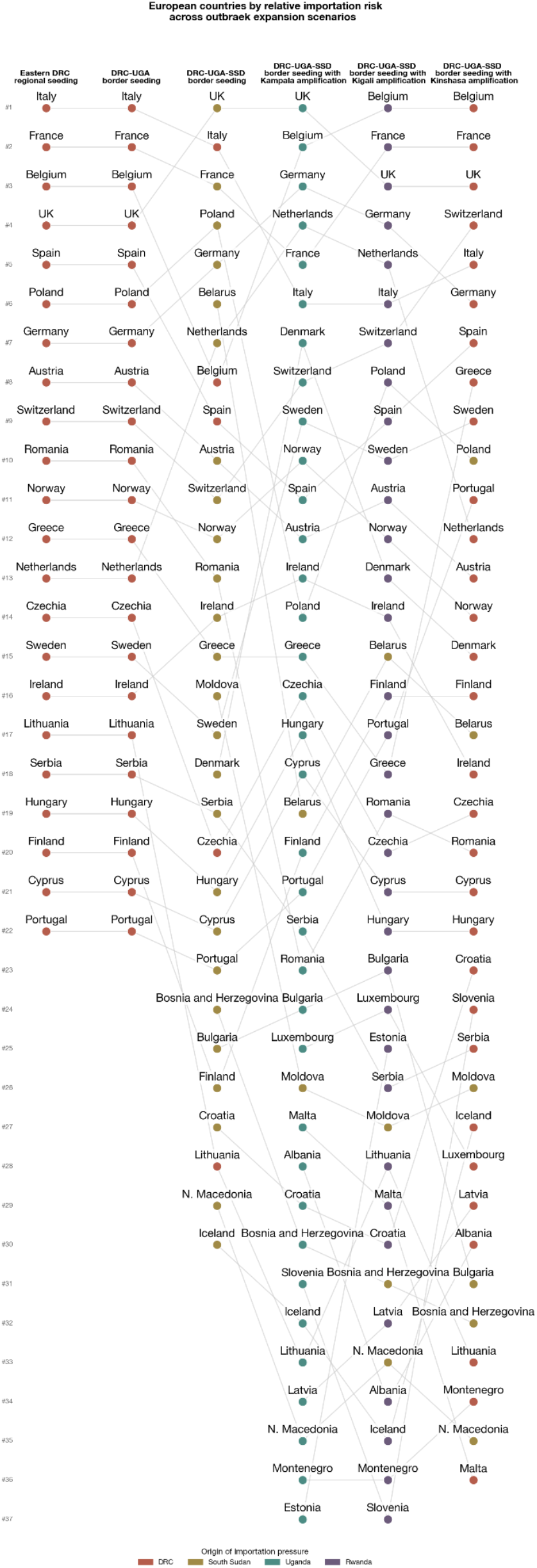
Ebola virus importation risk to Europe. Ranking of European countries by relative importation risk across outbreak expansion scenarios. Dot colors indicate the dominant origin of importation pressure (DRC, Uganda, South Sudan, or Rwanda). Analyses performed using average monthly travel data from the full 2024.

**Figure S3.**
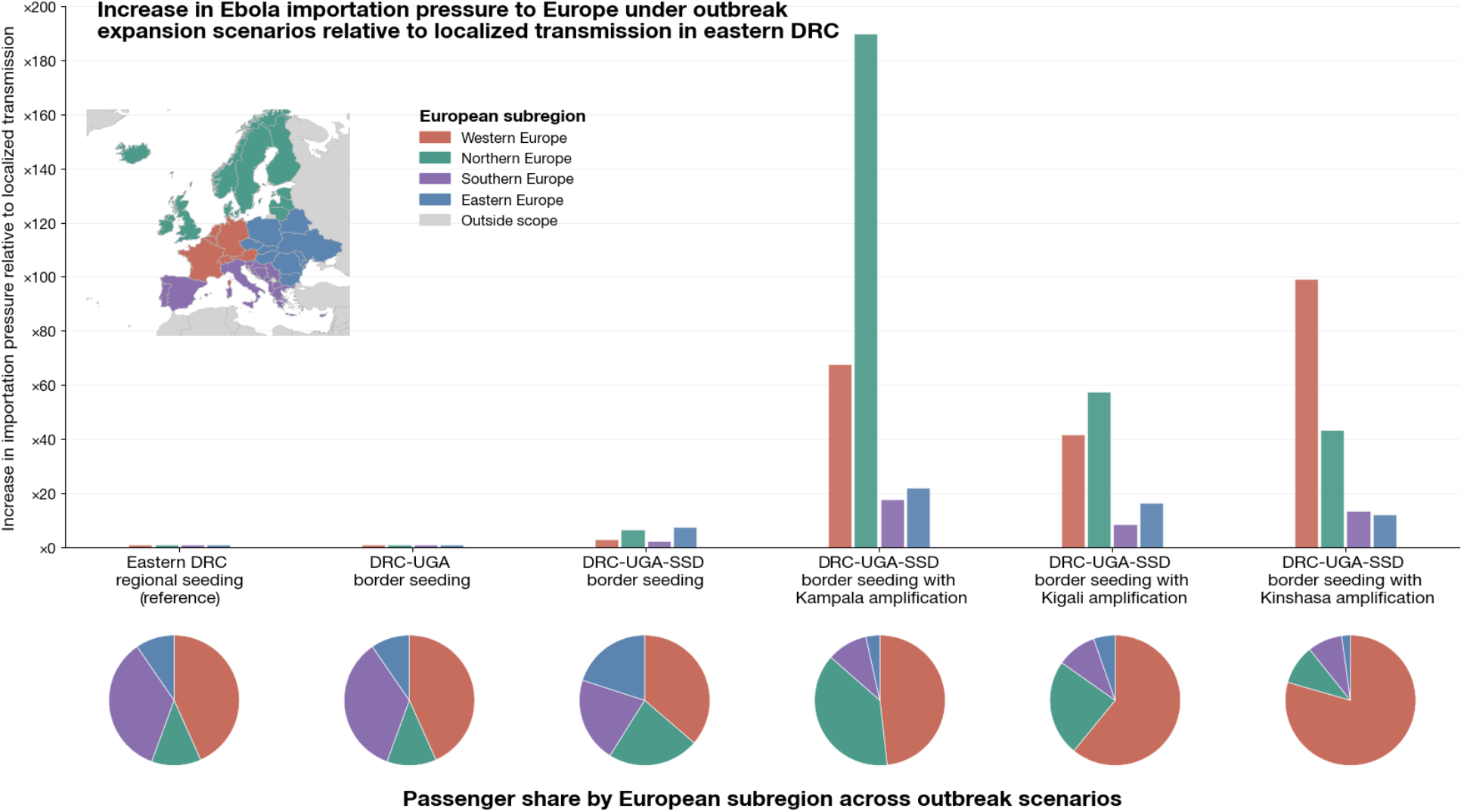
Increase in Ebola importation pressure in Europe. Increase in Ebola importation pressure to European subregions across outbreak expansion scenarios, compared with localized transmission in eastern DRC (reference scenario). Pie charts indicate the distribution of passenger flows across subregions for each scenario. Analyses performed using average monthly travel data from the full 2024.

**Table S1.**
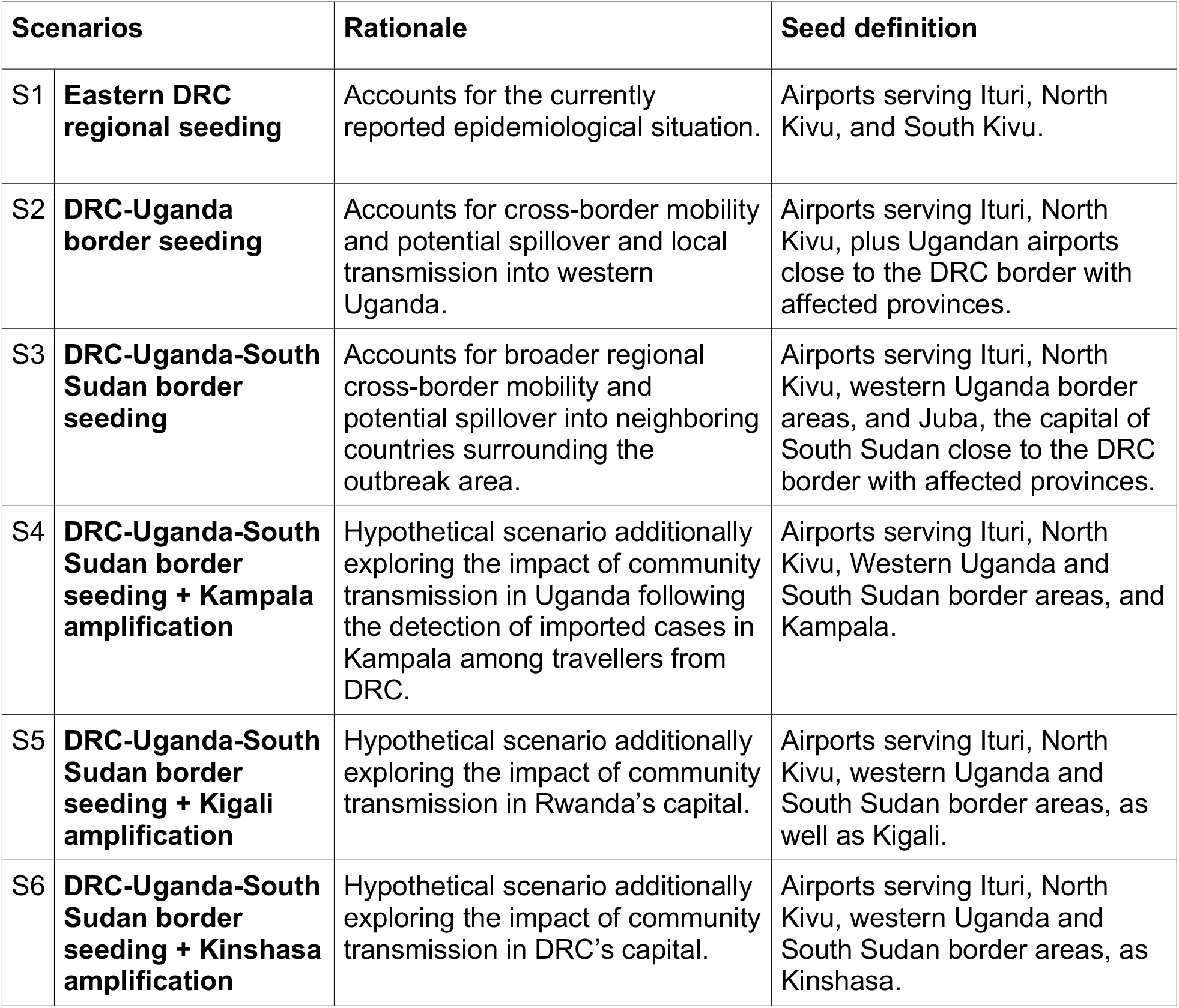
Outbreak expansion scenarios and source airports.

| Scenarios |  | Rationale | Seed definition |
| --- | --- | --- | --- |
| S1 | **Eastern DRC regional seeding** | Accounts for the currently reported epidemiological situation. | Airports serving Ituri, North Kivu, and South Kivu. |
| S2 | **DRC-Uganda border seeding** | Accounts for cross-border mobility and potential spillover and local transmission into western Uganda. | Airports serving Ituri, North Kivu, plus Ugandan airports close to the DRC border with affected provinces. |
| S3 | **DRC-Uganda-South Sudan border seeding** | Accounts for broader regional cross-border mobility and potential spillover into neighboring countries surrounding the outbreak area. | Airports serving Ituri, North Kivu, western Uganda border areas, and Juba, the capital of South Sudan close to the DRC border with affected provinces. |
| S4 | **DRC-Uganda-South Sudan border seeding + Kampala amplification** | Hypothetical scenario additionally exploring the impact of community transmission in Uganda following the detection of imported cases in Kampala among travellers from DRC. | Airports serving Ituri, North Kivu, Western Uganda and South Sudan border areas, and Kampala. |
| S5 | **DRC-Uganda-South Sudan border seeding + Kigali amplification** | Hypothetical scenario additionally exploring the impact of community transmission in Rwanda’s capital. | Airports serving Ituri, North Kivu, western Uganda and South Sudan border areas, as well as Kigali. |
| S6 | **DRC-Uganda-South Sudan border seeding + Kinshasa amplification** | Hypothetical scenario additionally exploring the impact of community transmission in DRC’s capital. | Airports serving Ituri, North Kivu, western Uganda and South Sudan border areas, as Kinshasa. |

